# A machine-learning Approach for Stress Detection Using Wearable Sensors in Free-living Environments

**DOI:** 10.1101/2024.04.27.24305829

**Authors:** Mohamed Abd Al-Alim, Roaa Mubarak, Nancy M. Salem, Ibrahim Sadek

## Abstract

Stress is a psychological condition due to the body’s response to a challenging situation. If a person is exposed to prolonged periods and various forms of stress, their physical and mental health can be negatively affected, leading to chronic health problems. It is important to detect stress in its initial stages to prevent psychological and physical stress-related issues. Thus, there must be alternative and effective solutions for spontaneous stress monitoring. Wearable sensors are one of the most prominent solutions, given their capacity to collect data continuously in real-time. Wearable sensors, among others, have been widely used to bridge existing gaps in stress monitoring thanks to their non-intrusive nature. Besides, they can continuously monitor vital signs, e.g., heart rate and activity. Yet, most existing works have focused on data acquired in controlled settings. To this end, our study aims to propose a machine learning-based approach for detecting the onsets of stress in a free-living environment using wearable sensors. The authors utilized the SWEET dataset collected from 240 subjects via electrocardiography (ECG), skin temperature (ST), and skin conductance (SC). In this work, four machine learning models were tested on this data set consisting of 240 subjects, namely K-Nearest Neighbors (KNN), Support vector classification (SVC), Decision Tree (DT), and Random Forest (RF). These models were trained and tested on four data scenarios. The K-Nearest Neighbor (KNN) model had the highest accuracy of 98%, while the other models also performed satisfactorily.

## 1. Introduction

Stress has become a widespread and influential factor in modern life, affecting individuals mentally and physically. Therefore, it is crucial to identify stress early and take appropriate action. Our research aims to identify stress using machine learning models effectively and before. The ability of machine learning to analyze patterns and complex relationships within data makes it a promising area for research.

In engineering and medicine, early stress detection presents unique challenges and opportunities. Advancements in technology have led to the creation of innovative tools and methods for collecting, analyzing, and determining stress parameters objectively. As a result of continuous research and development, wearables, physiological sensors, and biometric data analytics have emerged as key components in the engineering arsenal of stress detection. This study analyzes measurements obtained from wearable sensors by machine learning algorithms to create robust models capable of discriminating stress-related subtle patterns.

In medical terms, stress refers to the body’s reaction to a difficult situation, such as severe anxiety when receiving an important e-mail or a phone call at an inappropriate time. Clinically, it refers to a psychological state of fear, severe anxiety, or distress that leads to physical and mental health problems. The most common physical problems associated with stress include weight loss, vascular disease, headaches, sleep problems, impaired immune function, and heart disease. In addition, stress can affect mental health and cause severe anxiety and depression [1].

Stress is considered one of the ten most important social determinants of health differences. It detrimentally impacts both mental and physical well-being, exerting a negative influence on both societal dynamics and economic factors. Thus, organizations such as the *Occupational Safety and Health Administration*, the *World Health Organization*, and the *American Psychological Association* consistently raise awareness about stress’s negative effects [2]. According to the statistics of the American Institute of Stress, 80% of employees suffer from stress during their shift, and statistics also highlight student suicides’ between the ages of 15 and 29 related to stress [3]. In recent decades, the rise in traffic accidents and fatalities has been linked to an uptick in driver fatigue, tiredness, and psychological strain [4].

Early detection of stress is crucial in preventing stress-related psychological and health problems. In addition, the approach to stress detection should be comprehensive and consider the physiological indicators and environmental conditions surrounding the person. The precision of stress detection has significantly increased through the seamless integration of machine learning into medical practices and the pervasive incorporation of wearable sensors into individuals’ daily lives. In addition, this approach enables the identification of stress based on an individual’s surroundings, facilitating meeting individual needs.

Stress does not just affect a person’s mental and physical well-being; it affects the well-being of a nation’s population, which has an economic and social impact, so it is essential to recognize stress early on. In general, identifying the environment surrounding individuals exposed to stress besides their physiological signals allows researchers to treat stress in its early stages, reduce the risk of chronic health conditions, and the ability to improve the quality of life. Thus, developing reliable early intervention strategies for stress detection shall involve lifestyle adjustments, behavioral interventions, and traditional targeted therapy.

Traditionally, stress detection relies on textual information gathered through interviews and questionnaires [5, 6]. Still, these methods could be improved given their time-consuming nature and inherent biases, providing only isolated glimpses into patients’ well-being. Overcoming these limitations, the present research has adopted innovative approaches, e.g., wearable sensors [7] [8] [9] [10] [11] [12] [13] [14] [15] and the inclusion of audio and visual data [7] [8] for timely and comprehensive stress detection. Despite the widespread use of questionnaire methodologies in stress detection, as previously outlined and further elucidated later, they still need to be improved. A significant drawback is an inherent inaccuracy stemming from self-reporting, which is susceptible to biases. Responses may be influenced by social desirability or a tendency to present oneself positively, revealing a clear element of self-bias. Furthermore, relying on self-reported stress levels raises concerns about individuals’ capacity to recall and articulate the extent of their stress accurately and consistently.

Additionally, the one-time nature of questionnaires may not capture the dynamic and evolving nature of stress since stressors and coping mechanisms can change over time. A questionnaire-based approach may not fully explore the nuanced aspects of stress individuals experience due to relying on predefined questions—the structured format of the questionnaire risks limiting individuals’ ability to express difficult aspects of stress. Further examination is needed to understand stress more deeply from the lived experience perspective rather than predefined indicators. Due to the previously mentioned gaps and shortcomings of questionnaire-based approaches, researchers should refer to physiological signals in addition to questionnaires as a complementary step.

Machine learning has brought about a revolutionary shift in stress detection, allowing researchers to avoid the pitfalls of earlier methods. It introduces powerful tools for efficiently processing and analyzing extensive datasets characterized by complex patterns. A paramount achievement of machine learning is its ability to grapple with the intricacies of stress-related phenomena, steering clear of the surface-level perspectives that traditional methods often succumb to. The strength of machine learning algorithms lies in their ability to recognize subtle correlations and distinctions in collected data and their resilience against the influence of social preferences. It is worth mentioning that fine-tuning these methods can continuously improve their performance and adaptability to individual variations and progressively increase their stress detection accuracy.

Various studies have been implemented to detect stress using physiological parameters. The reason is that many psychological factors can cause stress, such as persistent worry about losing a job, approaching a deadline, etc. These physiological changes result in the body’s “fight-or-flight” response [10]. The rates of specific vital signs can increase or decrease as a response to stress, such as an increase in the number of heartbeats or a decrease in body temperature. These changes in vital signs mirror a person’s physiological parameters, enabling researchers to detect stress using wearable sensors. This paper endeavors to identify physiological alterations induced by stress. The complex interaction between stress and the sympathetic nervous system releases cortisol and adrenaline, triggering a cascade of physiological responses. This hormonal release manifests in an elevated heartbeat, influencing respiratory patterns, inducing perspiration, altering body temperature, and causing muscle tension. Consequently, by vigilantly monitoring these nuanced physiological changes, the machine learning model gains the ability to effectively discern and detect the presence of stress [10] [13] [14]. Smartphones and smartwatches have been widely used in recent years. This technological revolution has given way to so-called wearable sensors that measure vital signs in a nailed way throughout the day. Wearable sensors enable continuous monitoring of vital signs and daily activities via electrocardiogram (ECG), respiration (RSP), blood volume pulse (BVP), Skin conductance (SC), and skin temperature (ST) sensors. Wearable sensor technology has surpassed its counterpart methods (e.g., questionnaires) in detecting stress thanks to its ability to monitor vitals continuously without disturbing daily life routines [16] [17].

This research introduces a machine learning-based approach for the early detection of stress. One important foundation for this research is a holistic view of stress, i.e., not looking at stress from one angle. This rule has been established to contribute to the development of effective stress detection models. Based on this rule, it was imperative to find a dataset containing physiological signals of stress, provided that these signals were collected under normal life conditions and not collected under controlled settings. The following section reviews the previous literature in the stress detection field. This review encompasses the implementation of machine learning models, a thorough exploration of dataset characteristics, and a presentation of the resultant findings. This interdisciplinary exploration aims to deepen the collective understanding of stress and various stress detection models, thereby contributing to overall well-being.

## 2. Related work

Research on stress detection using machine learning is rich and diverse, as different methodologies and approaches are explored to meet the complex challenges associated with identifying stress indicators. This section reviews some of the previous literature in the field of stress detection, summarizing the methodologies of these studies, the data sets used, and the main results. Given the pivotal role of machine learning models in stress detection research, this role should have been strongly highlighted. Support vector machines (SVMs), decision trees (DT), random forests (RF), and neural networks (NN) are just a few of the algorithms that have been used to analyze various datasets of physiological signals [18]. Diverse patterns that signify stress can be captured differently by these models.

Attempts to automate and improve stress detection have become increasingly common in recent years. At the same time, coordinated efforts have been made to create stress datasets to promote a deep understanding of stress and its management. While a limited number of stress datasets are publicly accessible, most remain proprietary and owned by dedicated research groups. It is necessary to carefully extract and curate relevant features to improve the accuracy of machine learning models in stress detection. A wide range of features are covered in this study, from behavioral characteristics closely associated with user interactions to time and frequency domain attributes obtained from physiological signals.

An essential part of stress detection is extracting frequency domain features from physiological signals. Sliding-time windows are widely used for removing time domain features from time series data segments [19] [20]. In [19] [21], frequency domain features are derived by focusing on high and low-frequency zones. Following this, essential statistical metrics such as kurtosis, mean, and standard deviation are computed, constituting a robust methodology for stress detection.

In the study by [22], emphasis was placed on datasets obtained from an array of physiological sensors encompassing electrooculogram, blood volume pulse, respiration rate, magnetoencephalography, acceleration, pulse oximetry, skin temperature, eye tracker, and ECG sensors. Forty-four physiological features were extracted to detect the stress via the eXtreme Gradient Boosting (XG Boost) technique. This study explored two datasets, including Snake and Cogload, each comprising 23 subjects. Notably, the authors achieved an accuracy rate of 80% for stress detection.

The study in [23] included a comprehensive analysis of classification and regression methods for stress detection. The researchers based their theoretical investigation on the AffectiveROAD dataset, a publicly available dataset compiled via an Empatica E4 sensor. The authors employed a Bagged tree-based ensemble for regression, while a random forest classifier was used for classification purposes. It is worth mentioning that the features derived from skin temperature and pulse blood volume sensors provided the highest accuracy, reaching 82.3% for regression and 74.1% for classification, respectively.

The study presented in [24] examined the effects of stress on adolescents. To explore this, the authors conducted research involving high school students, generating a dataset using the Empatica E4 wearable sensor worn as a wristband. The study involved eight students whose vital signs were monitored for four weeks. From this continuous monitoring, 756 features were extracted across various biomarkers. The classification process employed a random forest classifier, resulting in a remarkable accuracy rate of 89.4%.

In [25], the authors examined the relative sensitivity and specificity of common vital indicators of stress gathered from healthy individuals subjected to various induced emotional states. The most common biomarkers of stress detection (e.g., heart rate, skin conductance, heart rate variability, respiratory rate, respiratory rate interval, and muscle activation) were evaluated using the WESAD dataset. Five features were extracted from the vital indicators, and then the classification was carried out using logistic regression, with the highest accuracy reaching 85.71%.

In [26], the researchers utilized the WESAD dataset, accumulating 14 relevant features. A neural network was subsequently employed for classification, yielding an accuracy of 85%. In this context, a novel approach to acute stress detection focusing on electrodermal activity was presented in [27]. The authors carefully assembled their data set by collecting data from 75 volunteers using a single sensor. The stress detection accuracy was 94.62% and was achieved by extracting 14 distinctive features and employing classification through Random Forest (RF) and Support Vector Machine (SVM) classifiers.

One of the most famous and well-known stress detection datasets is WESAD [28], introduced in [10]. Despite its considerable recognition, the dataset comes from a relatively limited number of 15 subjects. The dataset is collected by attaching sensors to participants’ chests and wrists. It includes three-axis acceleration, electrocardiogram, blood volume pulse, body temperature, respiration, electromyogram, and electrodermal activity recorded by RespiBAN Professional and Empatica E4 [10]. This dataset classified stress into two classes: a binary representation (stress or no stress) and three levels (stress, baseline, amusement). Consequently, the highest accuracies for these classifications were 93.12% and 80.34%, respectively.

In a recent development, [29] introduced the SWELL-KW dataset, which is publicly accessible. This dataset originates from 25 volunteers engaged in office-related tasks such as reading and writing. The volunteers operated under two distinct working conditions: receiving emails and facing time constraints. Recorded data encompassed a spectrum of parameters, including facial expressions, skin conductance, body positions, computer usage, and heart rate. This dataset was evaluated through questionnaires rigorously scrutinized for their effectiveness and alignment with mental effort and task load measures.

Furthermore, a large-scale cross-sectional study, a.k.a. SWEET, to detect stress included 1002 participants [30]. Throughout the study, vital signs were collected for each volunteer through smartphones and wearable devices over five consecutive days. Two wearable devices were used to monitor skin temperature, electrocardiogram, and skin conductance. The first of the two devices is the chest patch, and it was designed to measure the electrocardiogram and acceleration at a sampling rate of 256 and 32 Hz. The second device was the imec’s Chillband worn on the wrist to monitor skin temperature, skin conductance, and acceleration sampled at 256, 1, and 32 Hz, respectively. As for the device worn on the wrist, participants were advised to use it only during the day and at night and take it off during bath times and strong physical exertion. For the chest patch, participants were advised to wear it day and night during normal daily life tasks such as bathing. Participants were warned to take it off during strong physical exertion.

While there has been a significant advancement in machine learning for stress detection, numerous obstacles remain. The lack of a single, comprehensive data set, the variations in health and ethnicity among individuals, and the gender of the model—which is thought to be complex—are the main causes of these limitations. These challenges have caught our attention and motivated us to make a concerted effort to understand this complexity better. As far as our pursuit of a better life is concerned, our main goal and concern is the ability to detect stress with high accuracy. Therefore, we strongly believe in the need to train machine learning models on physiological signals and everyday activities. We chose a dataset collected while the volunteers were performing their daily lives because we aim to improve individuals’ lives to cope with stress.

Based on the findings from previous studies of relevant stress detection research, our methodology seeks to fill existing gaps and overcome challenges by using a range of machine learning models. Diverging from the controlled settings often employed in initial studies, our approach adopts a more ecological and lifelike context. The dataset used for this study was collected while subjects participated in their daily routine activities without consciously induced stressful scenarios. This approach envisions capturing the complexity and variations of stress in the free-living environment and promoting a more precise understanding of stress recognition in individuals’ daily lives.

## 3. Methodology

### 3.1. SWEET Dataset

This work used the dataset presented in [30] with prior permission. In [30], the study known as SWEET (stress in the work environment) was carried out on 1002 healthy adult volunteers (484 males and 451 females, in addition, of whom 76 volunteers did not fill out the questionnaires correctly) whose vital signs were monitored over five days a week (Thursday to Monday) for two years. The ages of the 1,002 volunteers ranged from 29.6 to 49.4. They work in 11 companies, some of which are in the public sector, some of which are banking and technology-oriented.

The volunteers in this study conducted four psychological questionnaires to assess the level of general health, anxiety, basic stress, sleep, and depression. Vital signs collected were ST, SC, and ECG via two portable devices. Both were used to measure the 3D ACC signal, which is used to control motion artifacts and estimate the intensity of physical activity. The first of the two devices is a chest patch intended for measuring the ECG and ACC with a sampling rate of 256 and 32 Hz, respectively. The second wearable device is the imec’s Chillband; this device is worn on the wrist and designed to measure SC, ST, and ACC and sampled at 256, 1, and 32 Hz, respectively.

The experiment participants were instructed to wear the bracelet during the day and remove it at night. The chest patch is worn during the day and night. Additionally, participants were asked to take off the bracelet when showering and both devices when doing intense physical activity. This dataset contained physiological data and periodic questions indicating the most important aspects of daily life activities and consumption. The authors [30] designed a smartphone app to alert participants to complete questionnaires.

The smartphone application alerts the participants in the experiment twelve times a day to fill out questionnaires randomly so that the duration between the two consecutive questionnaires is approximately 30 minutes. The questionnaire consists of four brief questions. The first question is about emotional feelings related to stress, namely dominance, pleasure, and arousal (i.e., level of control). The second question was about the stress level on the 5-point Likert scale, and stress was not asked by choosing between three options: low, medium, and high stress.

A person’s behavior in daily life when eating and drinking, as well as their regular daily activities, can affect the function of organs. The third and fourth questions were intended to cover this part of the life of the individual participating in this experiment. The planned consumption was mainly on food and drinks. Hence, the volunteers had to fill out a questionnaire with the type of drink or the name of the meal they ate by a particular number, as shown in Table 1.

**Table 1.**
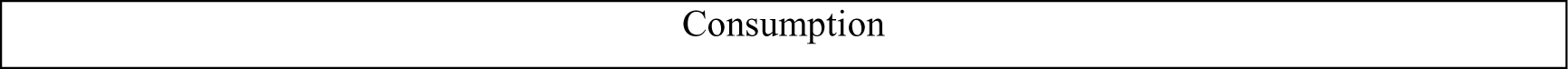
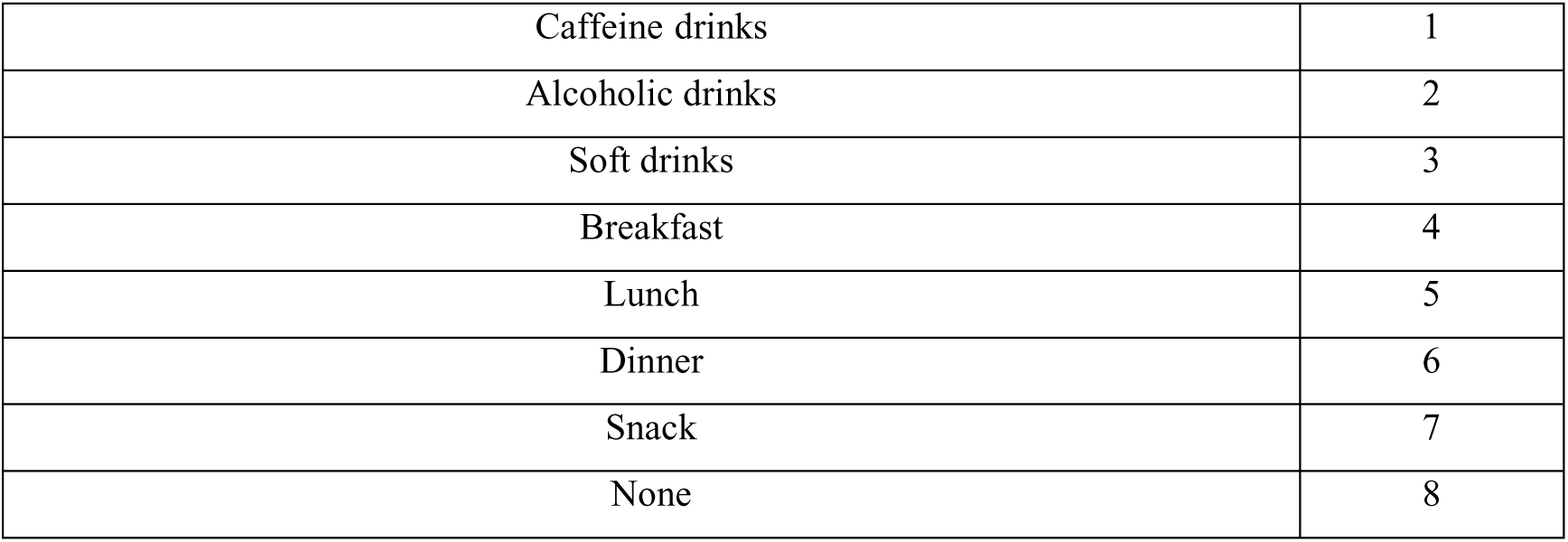
Overview of subject consumption.

|  |
| --- |
| Consumption |

|  |  |
| --- | --- |
| Caffeine drinks | 1 |
| Alcoholic drinks | 2 |
| Soft drinks | 3 |
| Breakfast | 4 |
| Lunch | 5 |
| Dinner | 6 |
| Snack | 7 |
| None | 8 |

Also, activities carried out by volunteers were included in the questionnaire, given that each activity has a particular number similar to the consumption (Table 2).

**Table 2.** Overview of subject activity.

| Activity |  |
| --- | --- |
| Lying down | 1 |
| Sitting | 2 |
| Standing | 3 |
| Walking | 4 |
| Running | 5 |
| Biking | 6 |
| Driving the car | 7 |
| Something else | 8 |

The datasets we made available contained 240 volunteers, with 5 days for each volunteer. It is worth mentioning that readings of the different sensors needed time-synchronization. As a result, a re-synchronization was performed to align readings across all sensors.

### 3.2. Pre-processing

Evaluating signal quality is critical because sensor readings can malfunction due to incorrect placement. As mentioned in [31], one of the ECG quality indicators is that the heart rate values should be limited between 40 and 180 BPM. In addition, the ECG readings are viewed from a different perspective, namely the accelerometer readings, so the ECG readings are acceptable when the standard deviation values are higher than 0.04. However, the ECG readings are discarded if the standard deviation is less than 0.04. After deleting ECG measurements with a standard deviation of less than 0.04, the remaining measurements should be continuous for ten minutes after deletion. If the measurements do not span a full ten-minute window, these measurements are deleted to ensure stability of the measurements.

Furthermore, according to [32], the values of SC should be higher than 0.001 µS and lower than 0.9 S. To maintain the stability of the readings output by the SC sensor, the first 15 minutes of readings are deleted due to irregular sweating immediately after wearing the band; In addition, due to the instability of the readings, the last 10 seconds of readings will be deleted before removing the bracelet. Additionally, readings under 3 seconds will be deleted, and the duration of correct readings should not be less than a 5-second window with 8 readings per second. The difference between the readings of one second should not exceed 20%, and the difference between them should not be less than 10%; otherwise, during one of the seconds, 8 readings will be omitted for that second. According to [33], one of the quality indicators for skin temperature is that the values are limited to 20 to 40 degrees Celsius.

Based on the above limits, outliers were removed. After combining all user data, we created 9655 samples for 238 users, each representing 1 minute of recording. The questionnaires from 238 volunteers on stress were distributed as follows: 71.7% had level 1 stress (6921 total samples), indicating no stress; 21% level two stress (full sample 2024); 5.2% level three stress (total 504 samples), 2.1% level four stress (total 204 samples), and almost 0% level five stress (total 2 samples), indicating the highest stress level, as shown in Figure 2.

**Figure 1.**
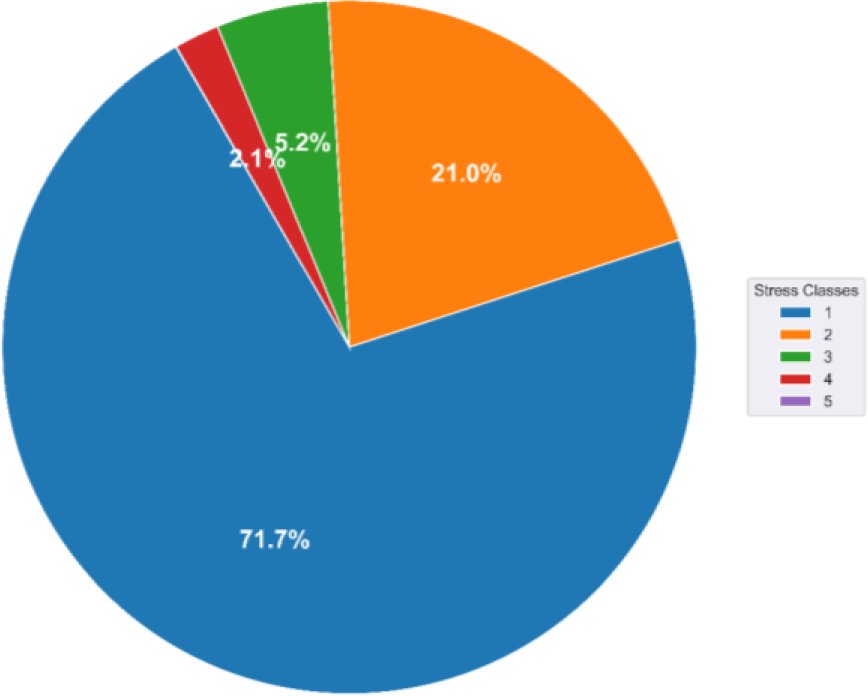
Distribution of samples in five stress levels.

**Figure 2.**
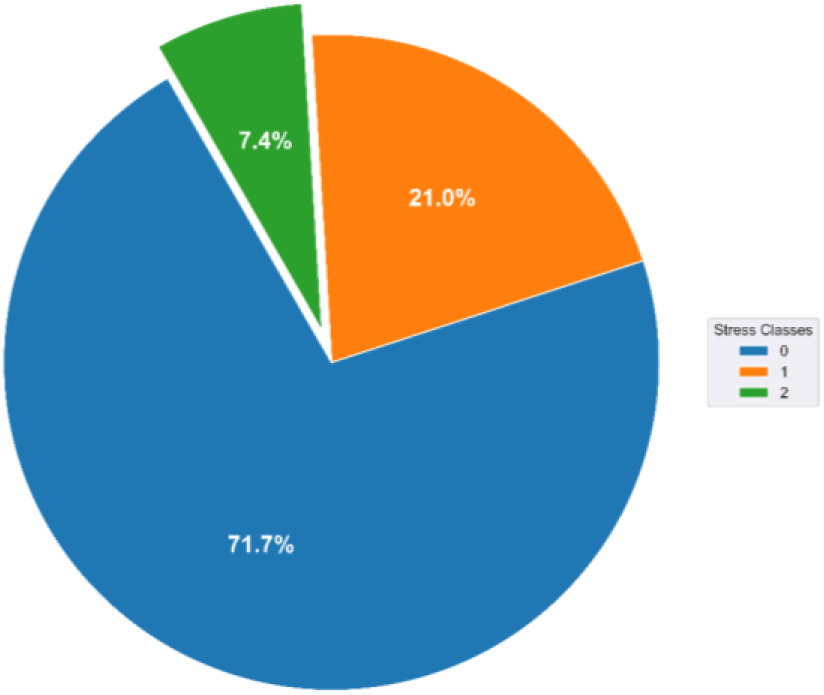
Distribution of samples in three stress levels.

Since our research aims to identify stress and its severity, we only looked at the times when the volunteers chose one of the five stress levels. It has already been pointed out that the authors [30] did not make any selection in the questionnaire regarding the state of stress in this form (no stress at all, mild, moderate, very tense). The authors rated the stress level in five levels, from 1 to 5. If volunteers choose No. 1, there is no stress; if they choose No. 5, it means they are under maximum stress.

The authors found Self-reported stress responses to be highly unbalanced [30]. For this reason, we suggest combining the three highest stress levels into one. Put another way, options 3 through 5 were merged into a single level, considering three stress levels as opposed to five (level 0 = no stress, level 1 = light stress, and level 2 = high stress). This results in the number of samples for Level 0 (6921 samples), Level 1 (2024 samples), and Level 2 (710 samples), representing 71.7%, 21%, and 7.4% of the total number of samples, respectively, as indicated in Figure 2.

After merging, the data was still unbalanced; thus, SMOTE (Synthetic Minority Oversampling Technique) was utilized to solve this issue. SMOTE is a resampling technique commonly used in machine learning to correct class imbalances in datasets, particularly in classification.

One of these problems is the bias of the classifier to the class containing a larger number of samples. After using SMOTE, the number of samples (6922) at each of the three stress levels is shown in Figure 3.

**Figure 3.**
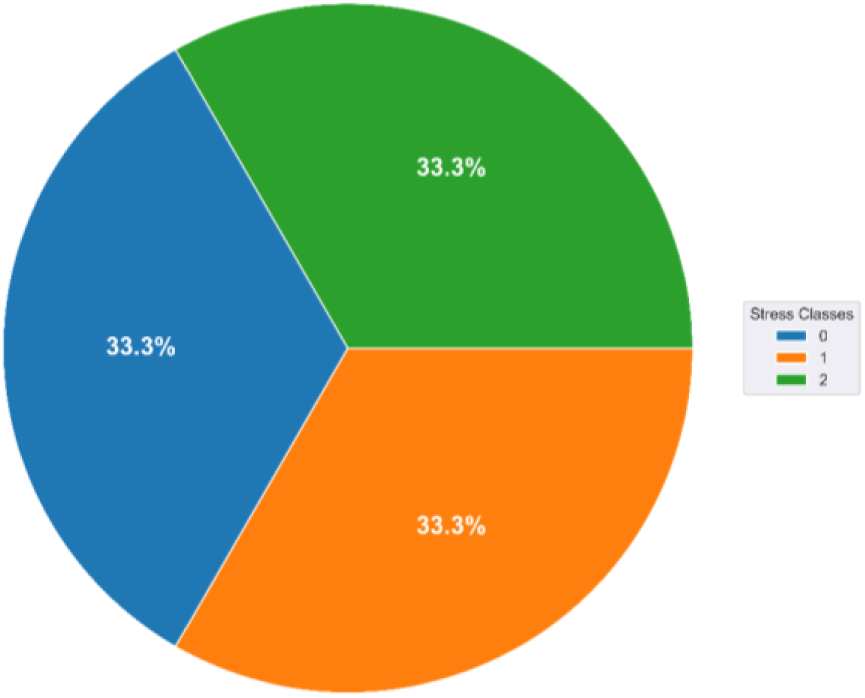
Distribution of samples into three stress levels after using the SMOTE technique.

Based on the approach presented in [30], we summarized the stress levels from level 2 to level 5 on the five-level stress scale into one level, called level 1, indicating the presence of stress. Furthermore, Level 1 on the five-point scale was maintained without transitioning to Level 0, again showing the absence of stress.

We then ended up with a binary classification problem where level 1 indicates the presence of stress, contains samples from levels 2 to 5 on the five-level stress scale, and level 0 indicates the absence of stress. It includes samples from level 1 of the five-level stress scale. The number of stage 0 samples (6921 samples) represents 71.7% of the total samples, in contrast to the number of stage 1 samples (2734 samples), representing 28.3% of the total samples, as shown in Figure 4.

**Figure 4.**
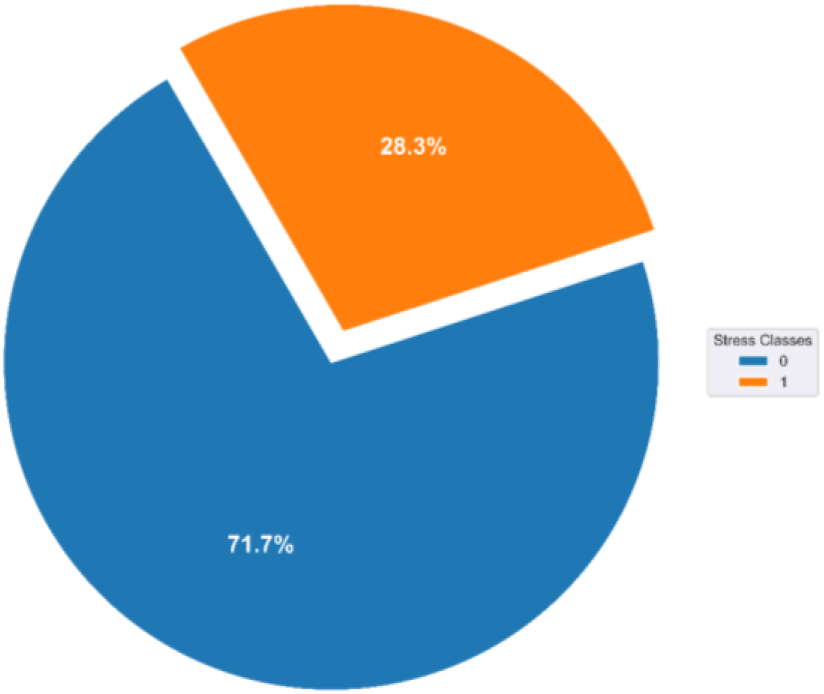
Distribution of samples into two stress levels.

It can be seen that even after converting the stress levels from 5 to only two levels, the problem of data imbalance still exists, which also required the use of SMOTE in this case. After using SMOTE, the number of samples in both layers was (6922 samples), as shown in Figure 5.

**Figure 5.**
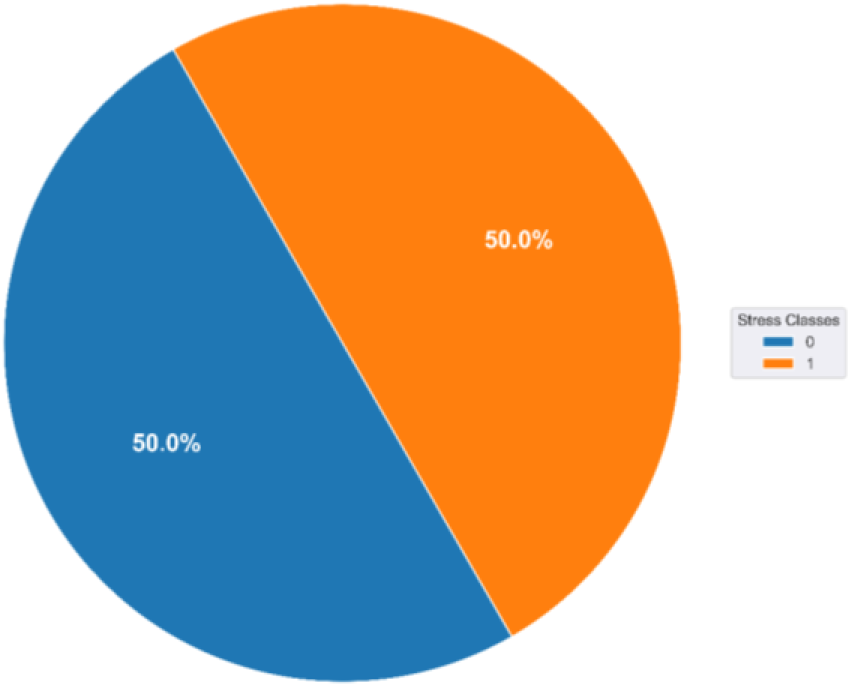
Distribution of samples into two stress levels after using SMOTE.

### 3.3. Feature extraction

There are several methods to extract features from physiological signals, including time-domain features, frequency-domain features, and statistical-based features. As mentioned in [34], statistical features such as min, max, mean, and standard deviation (std) have achieved good results in stress classification. By contrast, flatness and skew showed modest performance in this task. The authors in [30] calculated 19 features of physiological signals of good quality were divided as follows (6 features for ECG, 4 features for ST, 8 features for SC, and 1 feature for ACC); the description of these features is shown in Table 3.

**Table 3.** Calculated features and their descriptions.

| Nr. | Physiological signal | Feature | Abbreviation |
| --- | --- | --- | --- |
| 1 | ECG | 1. Mean heart rate (HR) | ECG mean HR |
| 2 |  | 2. Standard deviation of RR intervals | ECG SDNN |
| 3 |  | 3. Root mean square of successive RR differences | ECG RMSSD |
| 4 |  | 4. Low-frequency signal (power in the 0.04-0.015 Hz band) | ECG LF |
| 5 |  | 5. High-frequency signal (power in the 0.15-0.4 Hz band) | ECG HF |
| 6 |  | 6. Ratio of low and high frequency | ECG LFHF |
| 7 | SC | 1. SC level – average SC | SC mean |
| 8 |  | 2. Phasic SC – signal power of the phasic SC signal | SC phasic |
| 9 |  | 3. SC response rate – the number of SC responses (i.e., peaks) in the window divided by the total length of the window (i.e., responses per second) | SC RR |
| 10 |  | 4. SC second difference - signal power in second difference from the SC signal | SC diff2 |
| 11 |  | 5. SC response - number of SC responses | SC R |
| 12 |  | 6. SC magnitude - the sum of the magnitudes of SC responses | SC mag |
| 13 |  | 7. SC duration - the sum of the duration of SC responses in seconds | SC dur |
| 14 | | 8. SC area - the sum of the area of SC responses in seconds. The area is defined using the triangular method ($1/2 * SC \text{ mag} * SC \text{ dur}$) | SC area |
| 15 | ST | 1. Mean ST | ST mean |
| 16 |  | 2. Median ST | ST median |
| 17 |  | 3. Standard deviation ST | ST SD |
| 18 |  | 4. Slope of the ST – the slope of a straight line fitted through the data | ST slope |
| 19 | ACC | The standard deviation of the magnitude of the accelerometer signal – a measure of movement intensity | ACC SD |

The authors in [30] found that consumption of caffeinated drinks or breakfast corresponded to high levels of stress; on the contrary, during dinner or alcohol consumption, this corresponds to a low level of stress, and this has been demonstrated in our research through the correlation matrix shown in Figure (6). The matrix visually represents the interplay between stress and key factors, offering insights into the intricate connections among physiological responses, lifestyle activities, and stress patterns.

In [34][35][36], the high frequency (HF) component represents cardiac parasympathetic nerve activity during rest, and the LF component represents sympathetic nerve activity during stress. Thus, the low-frequency (LF) and the Low-Frequency to High-Frequency ratio (LFHF) components are expected to be higher during stress conditions and the HF component to be lower. In general, RMSSD has been reported to be more reliable than LFHF, in particular, because of the mechanical effects of respiration on HF power and the impact of prevailing heart rate on LF power. After considering the recommendations mentioned in [30], the 19 features were reduced to only 10 features using the Benjamini-Hochberg procedure, namely 2 for the SC (SC_area and SC_phasic), 2 for the ST (ST_median and ST_std), and 6 for the ECG (ECG mean HR, ECG SDNN, ECG RMSSD, ECG LF, ECG HF, and ECG QI_mean).

Numerous features have been identified in this section based on physiological signals and contextual data gathered from the data set referenced in [30]. This thorough approach allowed us to investigate the complex nature of stress. In the following section, these features will be employed to train and enable our machine-learning models to identify complex stress patterns in real-life scenarios.

### 3.4. Classification

Several machine learning algorithms have been used in stress detection. In this section, a brief overview of these algorithms is provided. In addition, the hyperparameters of each model will be discussed, as well as how they can be adjusted to achieve optimal results.

#### 3.4.1. K-Nearest Neighbors (KNN)

As a non-parametric method for classification and regression, K-Nearest Neighbors (KNN) is introduced. The KNN approach makes predictions based on the output variable’s k-nearest neighbors’ average value or majority class. Researchers highlighted KN’s simplicity and flexibility, emphasizing its ability to adapt to complex patterns in the data without making strong assumptions about the underlying distribution [37]. KNN is a useful algorithm commonly used for classification tasks, making it an effective tool for predicting stress levels. The proximity-based approach enables the model to recognize patterns and relationships in the data. It was also mentioned in [38] the importance of distance metrics such as Manhattan distance or Euclidean in determining the sameness between data points.

Regarding classification and regression tasks, KNN is a simple and intuitive algorithm. The central principle of KNN is to predict the class or value of a data point depending on the average value or majority class of its k nearest neighbors in the feature space. In classification mode, the KNN algorithm classifies a new instance by identifying the class that occurs most frequently among its k nearest neighbors. In regression mode, the expected value is the average of the purpose values of the k nearest neighbors. KNN assumes that similar samples in the feature space should have the same output values. This article used the KNN technique to classify stress levels once into two levels and then again into three. The hyperparameters of KNN are optimized using grid search to ensure the highest accuracy, i.e., H. for all cases (binary case with the use of SMOTE, binary case without use of SMOTE, three classes with the use of SMOTE, and three classes without use of SMOTE) and were as follows n_neighbors = 3, P = 1 and weights = distance.

#### 3.4.2. Support vector classification (SVC)

Support Vector Machines (SVM) are powerful supervised learning algorithms for classification and regression. SVC is a particular type of SVM designed for classification tasks; the primary aim of SVC is to meet a hyperplane that best individualizes the data into various classes while maximizing the border between classes. A hyperplane’s border is defined as the distance between it and the nearest data point of any class, and the optimal hyperplane maximizes this distance. They may be referring to Support Vector Regression SVR, another variant of SVM designed for regression tasks.

This paper used the SVC technique to classify stress levels once into two levels and again into three levels. The hyperparameters of SVC are tuned using grid search to ensure the highest accuracy, that’s for all cases (binary case with using SMOTE, binary case without using SMOTE, three classes with using SMOTE, and three classes without using SMOTE) and were as follows the regularization parameter (c) = 10, gamma = 1, and kernel = Radial Basis Function (RBF).

#### 3.4.3. Decision Tree (DT)

Decision trees are one of the most widely used machine learning algorithms for classification and regression tasks. Based on the value of the input features, the input space is recursively divided into subsets, creating a tree-like structure where each internal node represents a decision based on a feature, each branch represents the possible outcomes of that decision, and the leaf node represents the final output. From the previous explanation of DT parts, the key components were root nodes, internal nodes, branches, and leaves.

DT process begins with picking the feature that best branches the data into subsets, striving to maximize the similar class labels (homogeneity)within each subset. This process is called Node Splitting. Then, the previous procedure is repeated for each subset, establishing child nodes and further sectionalization of the data until a stopping criterion is matched; this process is called Recursive Partitioning. Typical stopping criteria include a peak in tree depth, a minimum number of samples per leaf, and a homogeneity threshold.

In classification tasks, leaf nodes represent class labels. A decision at every internal node is based on a feature’s value to assign the correct class label. This paper used the DT technique as a classifier to classify stress levels once into two levels and again into three levels. The hyperparameters of DT are tuned using grid search to ensure the highest accuracy for all cases (binary case with SMOTE, binary case without SMOTE, three classes with SMOTE, and three classes without SMOTE).

In the case of two classes with SMOTE, the hyper-parameters were Criterion = ‘entropy,’ the advantage of choosing the entropy criterion to measure the information earned achieved by splitting nodes. At each node, it is used to determine the best feature to split on. Entropy focuses on minimizing uncertainty in class labels. Maximum Depth = None: choose the maximum depth to ‘None’ and enable the tree to expand till each leaf node is pure (contains samples of only one class) or till the minimum sample leaf criterion is matched. When choosing the maximum depth to be ‘None,’ prevent overfitting by permitting the tree to adjust to the complexity of the data. Minimum Sample Leaf = 1: this lower value allows the tree to generate smaller leaf nodes, probably capturing finer patterns. Minimum Sample Split = 2: In a binary classification situation, a node will not divide if the number of samples is less than this value, promoting the overall generalization of the model. These hyper-parameters were the same as they were in the case of three classes with SMOTE.

In the case of two classes without using SMOTE, The hyper-parameters were largely the same as that in the case in which the SMOTE was used, except that the parameter of minimum Sample Split = 5: choose a higher value such as 5 to ensure that the tree constructs more robust decision nodes, this ensures that the model is not biased towards classes that have a larger number of samples. As in the case of three classes without using SMOTE, the hyperparameters were the same as in the case of three classes with SMOTE, except Maximum Depth = 20. This is to help control the complexity of the model, which may prevent overfitting and improve generalization.

#### 3.4.4. Random Forest (RF)

Random Forest is an ensemble learning technique that establishes multiple decision trees and amalgamates their outputs to improve predictive performance and minimize overfitting. Every decision tree within the forest undergoes training on a randomly selected subset of the dataset. The ultimate prediction typically relies on a majority vote or averaging, contingent on the nature of the task, whether it involves classification or regression [37].

Random Forest employs bagging, generating numerous bootstrap samples (random samples with replacement) derived from the initial dataset. Each tree is then trained using one of these bootstrap samples. Each decision tree node randomly evaluates a subset of features for division. This introduces extra randomness and diversity among the trees, enhancing the overall resilience of the ensemble. The ultimate prediction is established in classification scenarios through a majority vote from the trees. In regression tasks, the typical approach involves calculating the average of the predictions made by each tree. The hyperparameters of RF are tuned using grid search to ensure the highest accuracy, that is for all cases (binary case with using SMOTE, binary case without using SMOTE, three classes with using SMOTE, and three classes without using SMOTE) and were as follows n_estimators=100 and random_state=42. The proposed machine learning models are trained over a private SWEET dataset [30]. One of the main factors that guided us to use the SWEET database is that it includes 1002 participants. However, only 240 were approved for the work, two of which were broken, so the task was done over 238 subjects. This number is significantly higher than the number of participants in other well-known stress detection datasets, like the SWAT database. Likewise, the other key factor in choosing this database is that the participants’ data were collected under normal life conditions, which allows us to have a more comprehensive understanding of stress.

The classification task became more challenging as a result of these two elements. This is caused by several things, one of which is that there will be an imbalance in the data because no laboratory intervention was used during data collection. Additionally, obtaining useful data from sensors when going about one’s regular business is very difficult because of the most basic everyday details, such as body movement and friction between the sensors and clothing. These factors all impact the sensors’ readings and can cause them to become noisy.

## 4. Results

Every machine learning model was evaluated using four different classification scenarios. In the first instance, the stress data was split into two unbalanced classes: those with and without stress. In the second scenario, we have the same material as the first two classes but have resolved the imbalance issue. The division of stress into three unequal classes—no stress, stress, and high stress—represents the third scenario. The fourth example is the same as the cases from the first three classes but with the imbalance issue resolved.

The problem of unbalanced data was solved using SMOTE. Employing the SMOTE, synthetic instances of the minority class are generated by interpolating existing samples. By doing this, the balance of the dataset is improved, and the model’s performance in underrepresented categories is improved. Thus, in the case of the binary without using smote, we get 2734 samples for stress and 6921 samples for no stress, and when using SMOTE, we have 6922 samples for both no stress and for stress equally. As for the case that we have three stress levels, without using SMOTE, we got 6921 samples for no stress, 2024 samples for medium stress, and 710 for high stress, and if we used SMOTE, we got 6922 samples for both no stress and for medium stress and high stress equally.

To assess the efficiency of the proposed method, we divided the existing samples into three parts and they are (training, validation, and testing). The samples were divided into training, validation, and testing to avoid problems such as overfitting, ensure the model adapts well to new data, and assess its effectiveness in real-world situations. This procedure assessed the trained model’s generalization ability to completely unknown individuals whose registrations were not included in the training data. To increase the efficiency of the models, the grid search function is used, which tunes better hyper-parameters.

The classification performance was assessed using accuracy, the F1-score, recall (sensitivity), precision (positive predictive values), and area under the curve (AUC). A method’s accuracy and AUC determine how well it performs across all classes of samples, whereas the F1-score, recall, and precision indicate how well it distinguishes between classes. Accuracy, precision, recall, and F1-score can be calculated as follows:

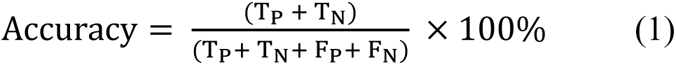

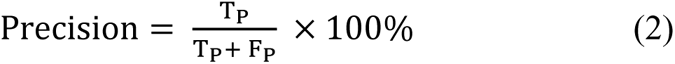

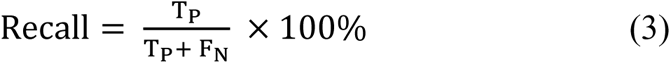

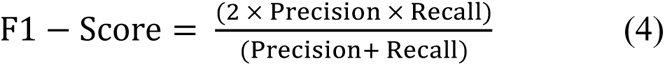

Where T_P_ is the number of correctly classified positive samples, F_P_ is the number of incorrectly classified negative samples. In this case, T_N_ is the number of incorrectly classified positive samples, and F_N_ represents the number of positive samples incorrectly classified as negative.

Accuracy (ACC) measures the classifier’s performance, quantifying the ratio between the correct classified classes and all predictions, as shown in equation (1). However, relying on the accuracy scale alone does not provide a complete picture of the model’s performance for two reasons. The first reason is that the accuracy scale is sensitive to data imbalance, an existing problem in our work that should be avoided. The second reason is that the model’s performance is very likely to differ from the performance of the other model. Still, their accuracy scale is the same because the accuracy scale in its calculation, as shown in equation (1), is based on the number of incorrect and correct predictions [39].

Consequently, recall and precision had to be used to fully capture the model’s performance from all angles and prevent models from becoming biased due to data imbalances. Equation (2) illustrates how the precision measure ascertains whether the dataset contains false positives, compromising overall accuracy. Equation (3) [40] describes recall, which measures how well a classifier predicts positive labels based on the quality of a match.

Confusion matrices help compare the classifier’s performance between the actual and predicted classes. The fundamental elements of the confusion matrix are True Positives (TP), True Negatives (TN), False Positives (FP), and False Negatives (FN). False Positives (FP) and False Negatives (FN) happen when the prediction does not match the actual emotional classes. When expected and actual emotional classes are positive and negative, respectively, TPs and TNs are applied. Although a negative class is anticipated, a positive class is seen. Both a positive and a negative predicted class apply to FN.

### 4.1. KNN Results

This section will review KNN’s performance and results through four data scenarios. In these four scenarios, the network search technique has adjusted the hyperparameters to ensure the best values are reached, which leads to the best results. The hyperparameters in the four cases were as follows: n_neighbors = 3, P = 1, and weights = distance. In Table 4, KNN’s performance during data handling for the four data cases will be detailed.

**Table 4.** Classification performance of the KNN model in four scenarios.

| Data scenarios | Accuracy | Recall | Precision | F1-Score |
| --- | --- | --- | --- | --- |
| Two levels without SMOTE | 98.447 | 97.994 | 98.203 | 98.097 |
| Two levels with SMOTE | 98.086 | 98.086 | 98.085 | 98.085 |
| Three levels without SMOTE | 98.343 | 97.840 | 97.752 | 97.793 |
| Three levels with SMOTE | 98.314 | 98.309 | 98.315 | 98.310 |

Looking at Table 4, two very important things become clear. The first is that the system becomes more stable when using SMOTE, as evidenced by the constant accuracy, recall, precision, and F1 score values. Second, the performance of KNN was similar in the four scenarios handled by KNN, and in [38], it was found that KNN can adapt to complex patterns in the data without making strong assumptions about the underlying distribution. For the picture to be complete and to evaluate the performance of the KNN machine learning classification model, the confusion matrix needed to be checked in four cases, as shown in Figure 7 (a, b, c, and d).

**Figure 6.**
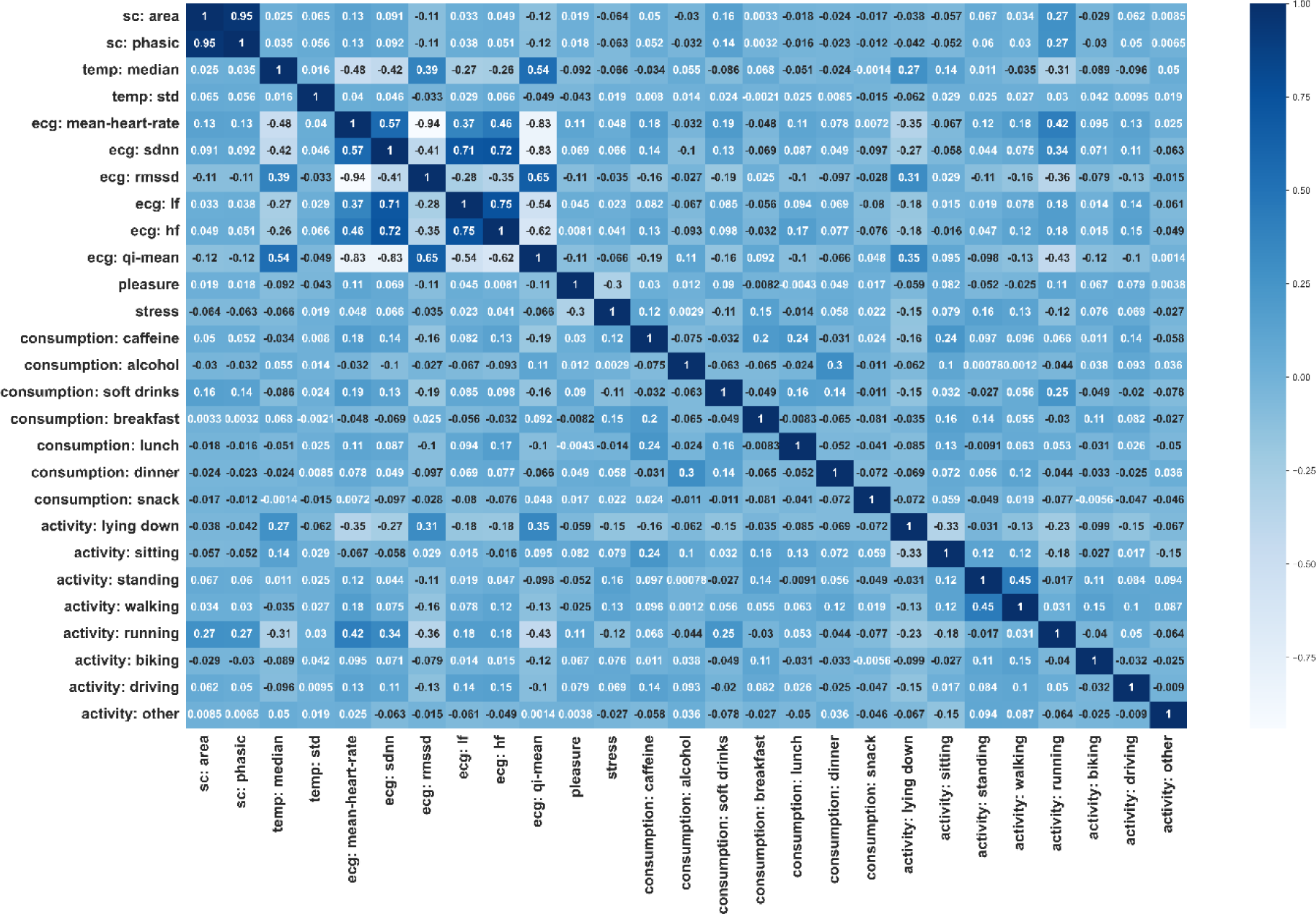
Correlation Matrix depicting the relationship between Stress and Physiological Signals features, Consumption, and Activities.

**Figure 7.**
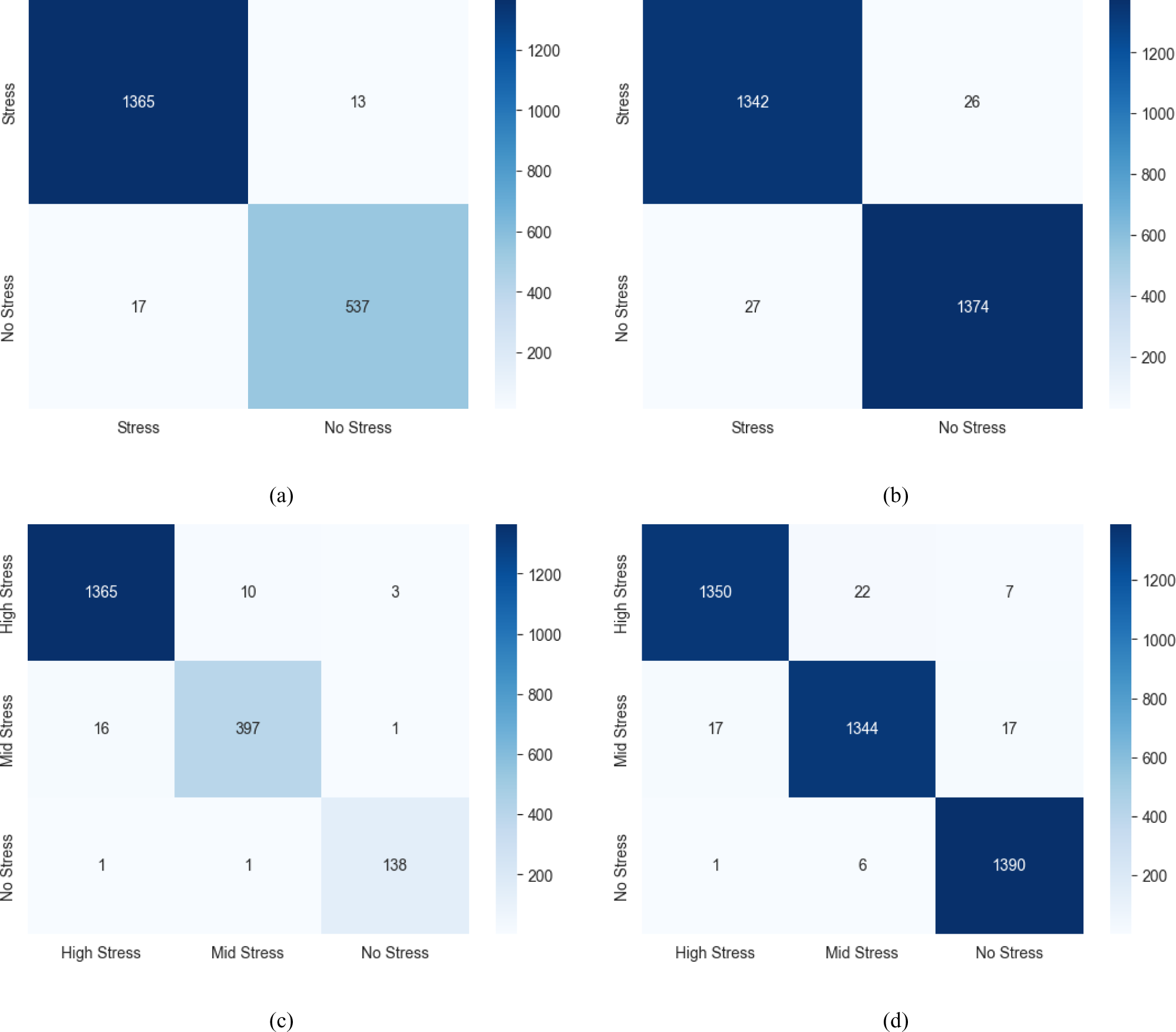
confusion matrix of four scenarios of the KNN model (a) the confusion matrix of two unbalanced classes, (b) the confusion matrix of two classes after using SMOTE, (c) the confusion matrix of unbalanced three classes, (d) the confusion matrix of three classes after using SMOTE.

In figure 7 (a, b, c, and d), the confusion matrix of the KNN machine learning model is checked for four cases. These are two levels of stress without using SMOTE, two levels of stress with using SMOTE, three levels without using SMOTE, and three levels without using SMOTE. From Figures (a, b), we can see that at only two stress levels, KNN could correctly classify 540 samples as stress states out of 553 samples selected to test the model. Furthermore, 1400 samples out of 1417 samples could be correctly classified. Figure 7(c and d) shows the model’s performance at three stress levels. Despite the small number of samples at this level, the model classified 140 out of 143 samples as highly stressed and only made no errors on three samples. After increasing the number of samples using SMOTE, KNN could distinguish 1400 samples as high-stress samples out of 1418.

### 4.2. SVC Results

SVC was tested in four scenarios, and its performance was examined separately. The model’s hyperparameters were adjusted using grid search, and the hyperparameters of the SVC were constant in the four scenarios where they were as follows: c = 10, gamma = 1, and kernel = RBF. In Table 5, the Accuracy, Recall, Precision, and F1-Score of the SVC’s performance in the four scenarios will be reviewed.

**Table 5.** Classification performance of the SVC model in four scenarios.

| Data scenarios | Accuracy | Recall | Precision | F1-Score |
| --- | --- | --- | --- | --- |
| Two levels without SMOTE | 96.273 | 94.311 | 96.528 | 95.336 |
| Two levels with SMOTE | 96.424 | 96.441 | 96.446 | 96.424 |
| Three levels without SMOTE | 95.807 | 92.803 | 96.501 | 94.539 |
| Three levels with SMOTE | 96.437 | 96.430 | 96.454 | 96.432 |

From Table 5, we can conclude that combining the five stress levels into just two levels helped significantly improve the performance of SVC in the absence of non-use of SMOTE. The second thing is that the smote did not stop its role in improving the performance of the SVC, but it made its performance more stable. To understand how SVC deals with the data in the four scenarios, it was necessary to display the confusion matrix, as shown in Figure 8.

**Figure 8.**
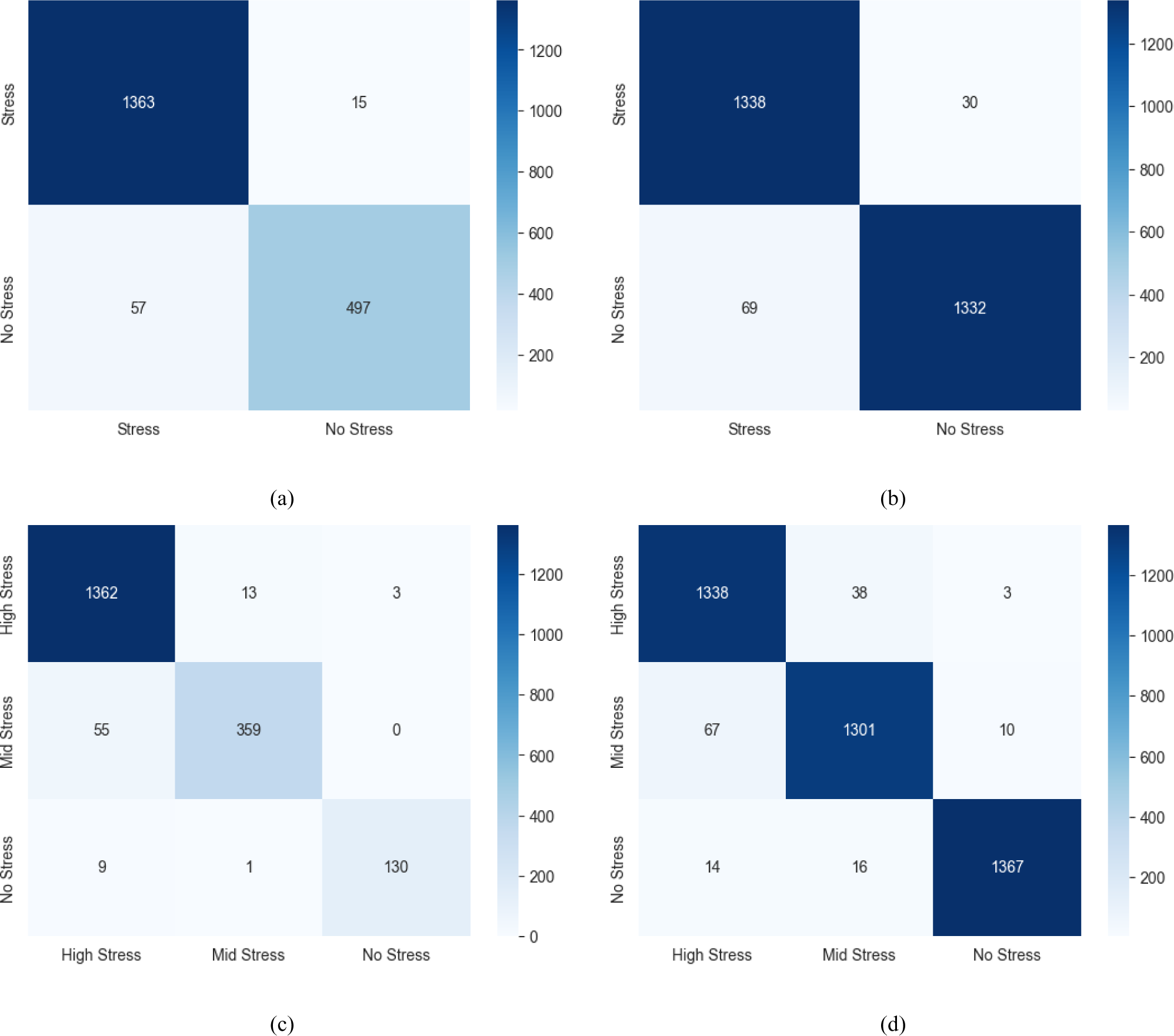
confusion matrix of four scenarios of the SVC model (a) the confusion matrix of two unbalanced classes, (b) the confusion matrix of two classes after using SMOTE, (c) the confusion matrix of unbalanced three classes, (d) the confusion matrix of three classes after using SMOTE.

Figure 8 clearly shows the effect of putting stress on only two classes in improving the efficiency of SVC. Using smote has also made SVC more stable and improved performance significantly.

### 4.3. DT Results

The model’s hyperparameters were adjusted using grid search, and the hyperparameters of the DT were not constant in the four scenarios and were summarized in Table 6 as follows. Table 7, Accuracy, Recall, Precision, and F1-Score, will review the DT’s performance in the four scenarios.

**Table 6.** DT models’ hyperparameters in four scenarios.

| Data scenarios | Criterion | Max_depth | Min_Samples_Leaf | Min_Samples_Split |
| --- | --- | --- | --- | --- |
| Two levels without SMOTE | Entropy | None | 1 | 5 |
| Two levels with SMOTE | Entropy | None | 1 | 2 |
| Three levels without SMOTE | Entropy | 20 | 1 | 2 |
| Three levels with SMOTE | Entropy | None | 1 | 2 |

**Table 7.**
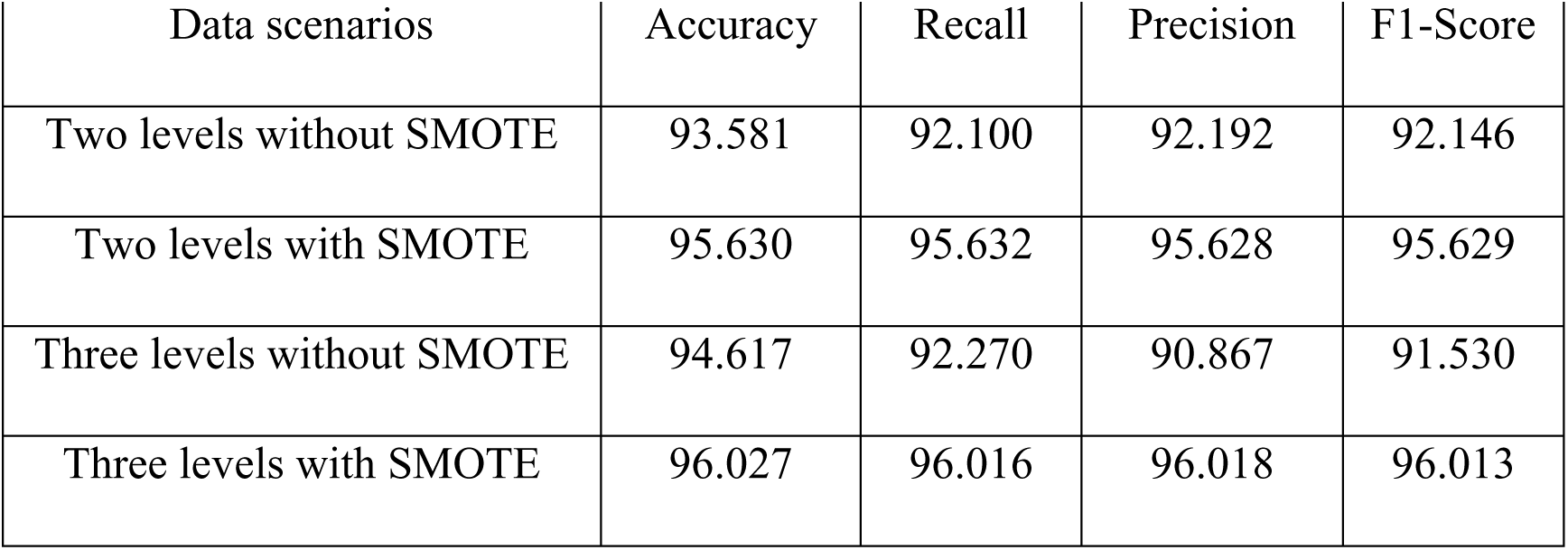
Classification performance of the DT model in four scenarios.

In Table 7, we review the DT’s performance in the four scenarios (i.e., Accuracy, Recall, Precision, and F1-Score).

Table 8 shows that splitting the data into two classes improves the model’s efficiency. This is clearly shown in the F1 score for both “Two Levels without SMOTE” and “Three Levels without SMOTE,” as the data imbalance exists in these two cases. Therefore, using the F1 score to measure the model’s performance is preferable. After using SMOTE, the model’s performance became more stable; it has also helped significantly improve its performance, as evidenced by the accuracy of all scenarios. In Figure (9), we review the confusion matrix for the four cases the model handled to show a complete picture of the model’s performance.

**Table 8.**
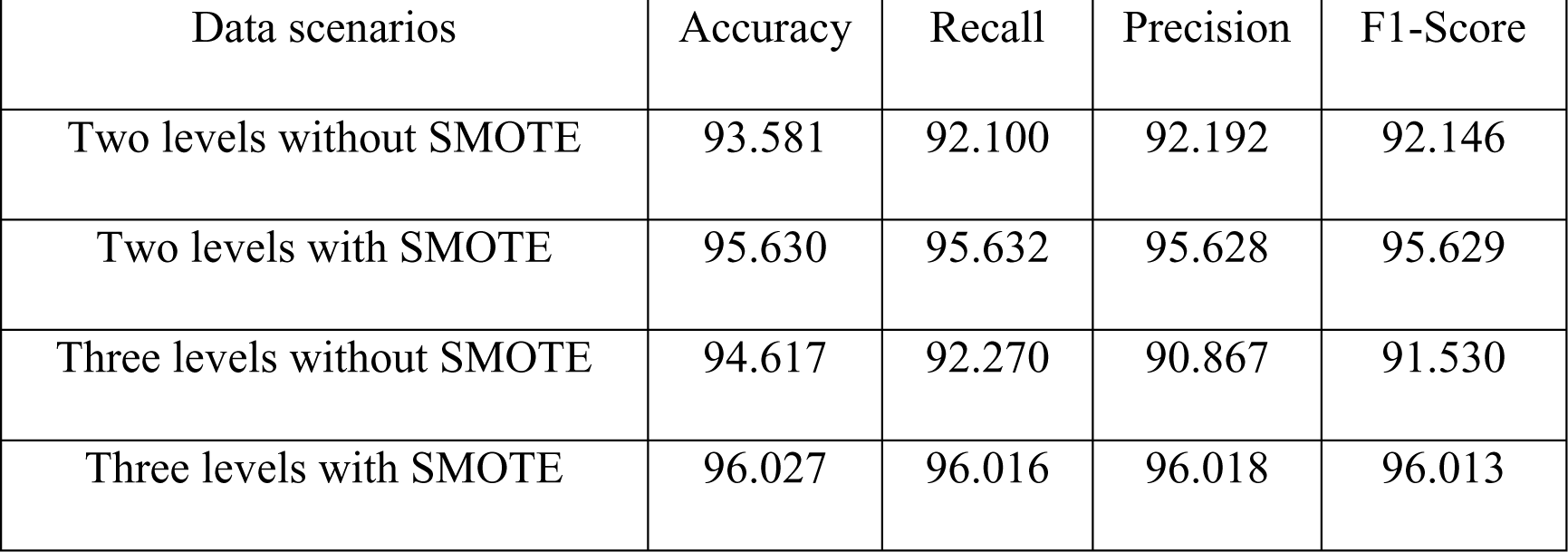
Classification performance of the RF model in four scenarios.

It is clear from Figure 9 that, as explained earlier, reducing the stress to only two classes instead of three enhances the model’s efficiency. In addition, it has also been made clear this time that using SMOTE improves the model’s efficiency and makes its decisions more stable.

**Figure 9.**
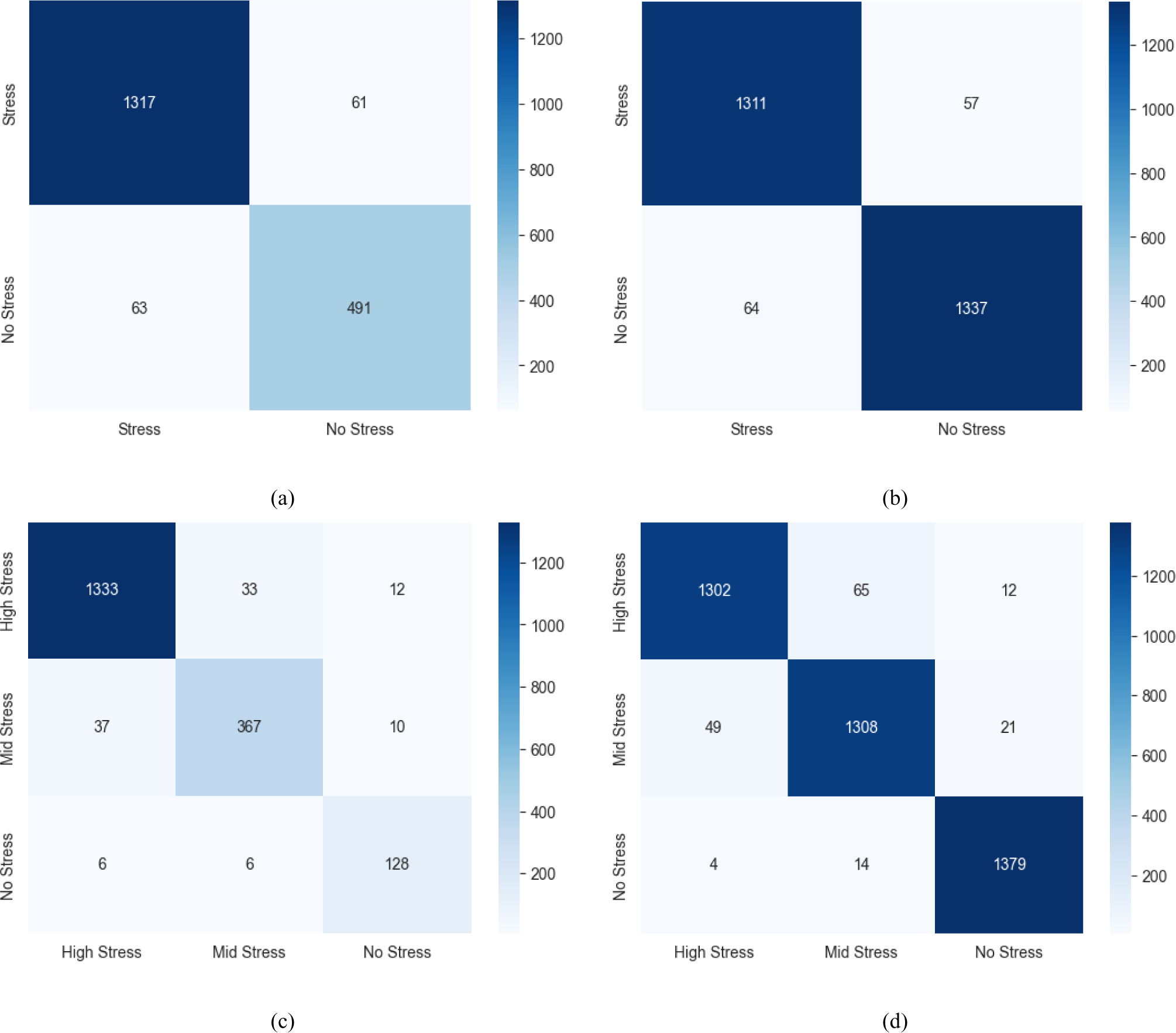
confusion matrix of four scenarios of the DT model (a) the confusion matrix of two unbalanced classes, (b) the confusion matrix of two classes after using SMOTE, (c) the confusion matrix of three unbalanced classes, (d) the confusion matrix of three classes after using SMOTE.

### 4.4. RF Results

DT’s work is very close to how RF works, but RF’s experience was important and necessary because RF was used in [30]. Because we also used the Grid search technique to tune the hyperparameters, it was challenging to train the model as it was introduced in [30] using a leave-one-subject-out approach, and here, the only difference was in the RF training method. The hyperparameters in the four data scenarios were n_estimators=100 and random_state=42. Table 8 provides the performance of the RF model with the four data scenarios.

Table 8 illustrates three key points. The first and second points are already known to us. Still, their recurrent appearance is interpreted as evidence of what is happening— that is, the model performs better when handling two stress classes than three. Further, when SMOTE is applied, the model’s performance is more stable and stronger. The third point is that RF performance is the same as DT performance, and that’s because they were trained in the same way. The best hyperparameters were selected for them in the same way, i.e., the similarity of the performance of the two models up to the matching. Below, we will consider the performance of the RF model more comprehensively by presenting the confusion matrix in Figure [10].

**Figure 10.**
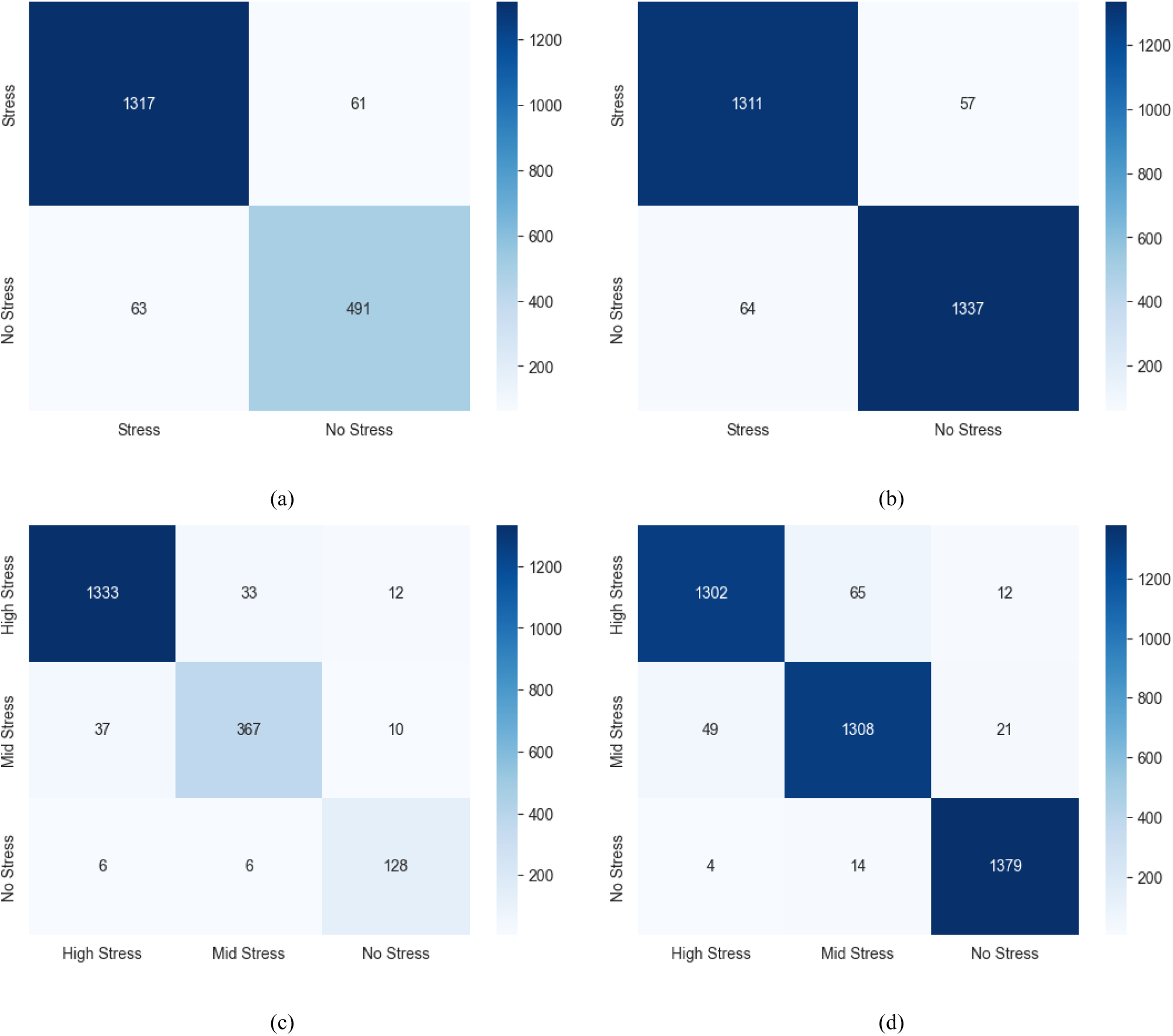
confusion matrix of four scenarios of the RF model (a) the confusion matrix of two unbalanced classes, (b) the confusion matrix of two classes after using SMOTE, (c) the confusion matrix of three unbalanced classes, (d) the confusion matrix of three classes after using SMOTE.

The performance of the four machine learning models was monitored in four different data scenarios.

It is clear from Table 9 that KNN was a superior performer in all four scenarios. Coming in second place is SVC, it had excellent performance to DT and RF in three scenarios (Two levels without SMOTE, Two levels with SMOTE, and Three levels without SMOTE), and its performance was almost the same as RF, and DT in the case of Three levels with SMOTE. As for the RF and DT performance, it was their best performance in the case of (Three levels without SMOTE and Three levels with SMOTE) they could deal with three levels of stress better than their ability to deal with two. In addition, in the two scenarios in which SMOTE was used, the values of Accuracy, Recall, Precision, and F1-Score were considered semi-constant for each of the models used in these two scenarios, which proves that the use of SMOTE helps stabilize the performance of the model. Figure 11 visually illustrates what is stated in Table 10, where Figure 11 (a, b, c, and d) shows the efficiency of each of the four models in Two levels without SMOTE, Two levels with SMOTE, Three levels without SMOTE, and Three levels with SMOTE, respectively.

**Figure 11.**
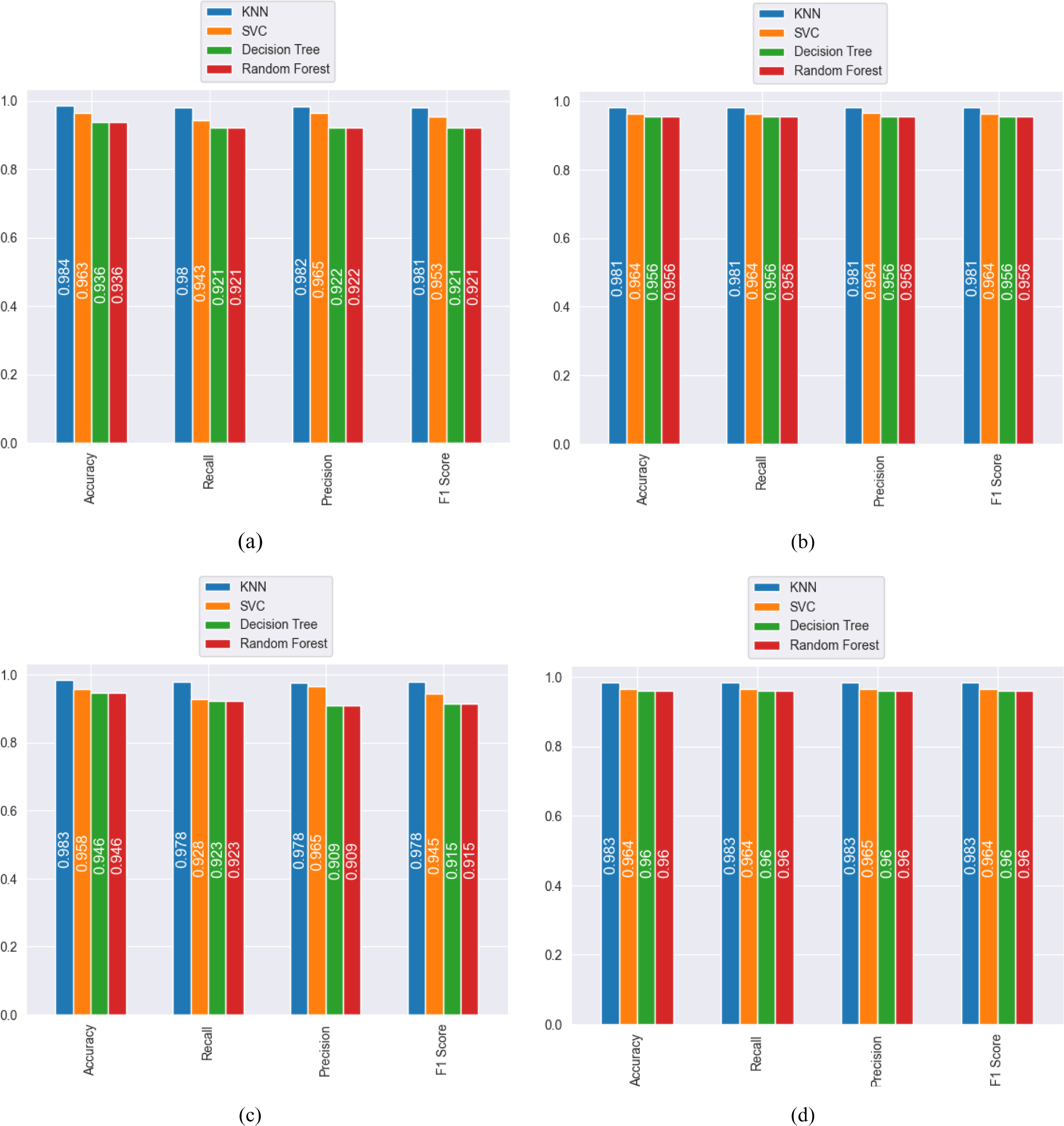
(a, b, c, and d) performance Comparison of KNN, SVC, DT, and RF in four scenarios: Two levels without SMOTE, Two levels with SMOTE, Three levels without SMOTE, and Three levels with SMOTE, respectively.

**Table 9.**
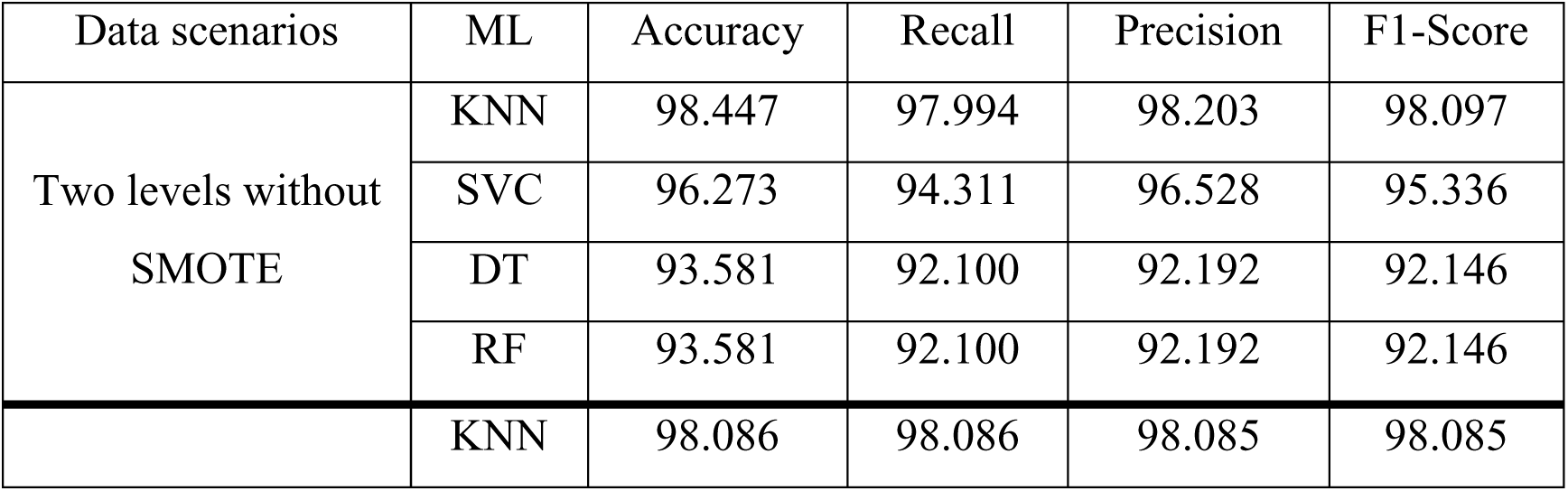
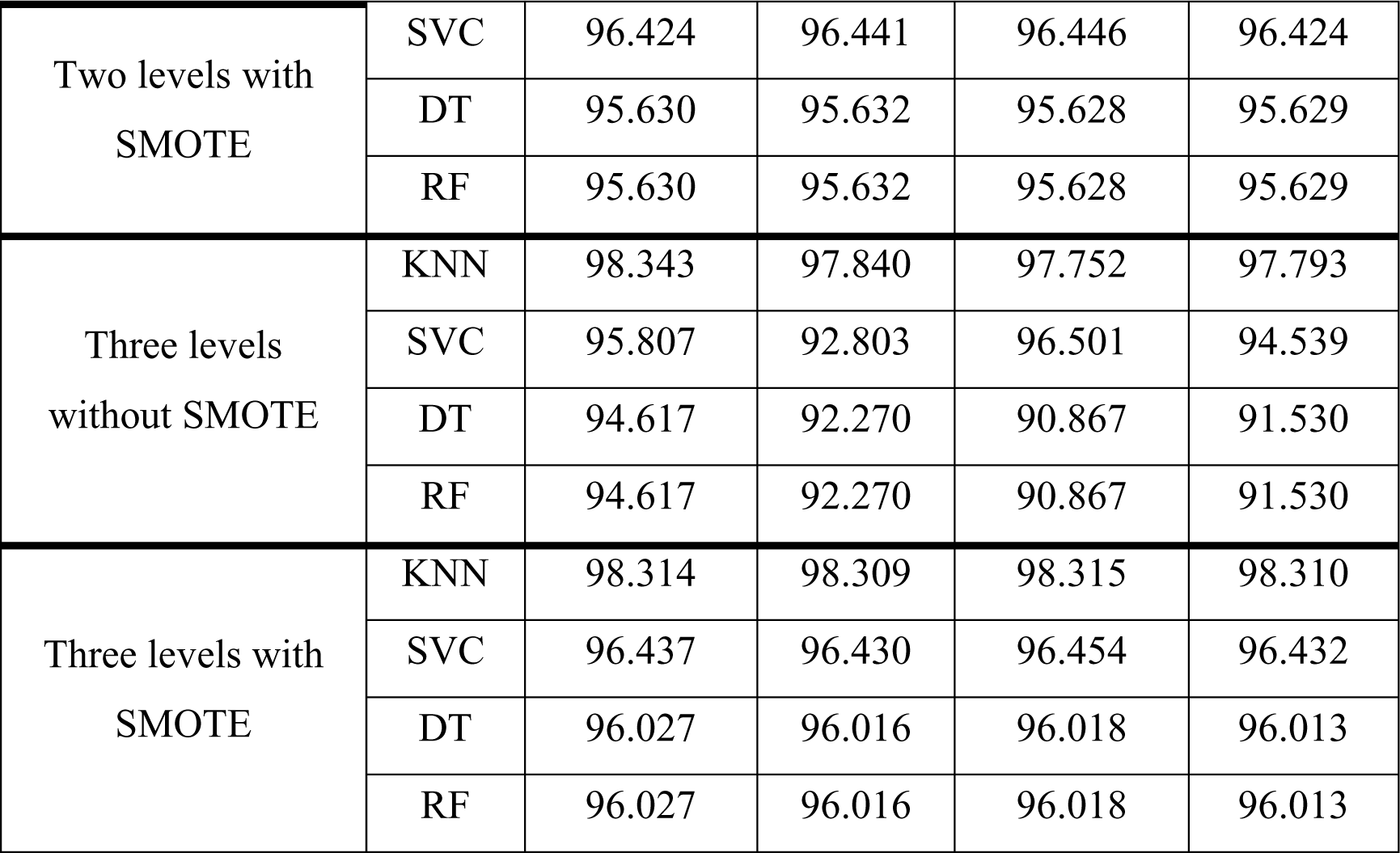
summarizes the overall classification results.

## 5. Conclusion

This study aims to go deeper into stress and look at stress from a broader perspective. Therefore, our quest was directed to using a realistic dataset, and the number of volunteers in it is large; for these reasons, the SWEET dataset was chosen. This database contains the data of 1002 subjects; 240 subjects were approved to use. This included two subjects whose data was missing, so work was done on only 238 subjects, a large number compared to the number of subjects in other databases that are widespread and widely used in stress detection. Since this database was collected under normal living conditions, it was natural that its data was unbalanced. The authors of the SWEET database divided stress into five levels. Then, the five levels were divided into only three levels, but these efforts did not help to solve the problem of data imbalance. In this work, an attempt was made to solve the problem of unbalanced data in two ways. The first way is to reduce stress levels to two levels instead of five; the other is to use SMOTE. Our exploration extended to the application of various machine learning models, namely K-Nearest Neighbors (KNN), Support Vector Classifier (SVC), Decision Trees (DT), and Random Forest (RF). Hyperparameter tuning, facilitated by the grid search technique, optimized the performance of each model. The four machine learning models were tested on four data scenarios. KNN has shown outstanding performance superior to SVC, DT, and RF. DT and RF performed similar performances to the point of unity since they are from the same family and work similarly. In conclusion, our study contributes novel insights into stress detection using real-life datasets and pioneers innovative strategies for handling imbalanced data. Integrating binary stress classification and SMOTE opens opportunities for further refinement of stress detection methods. A detailed analysis of machine learning models reveals their unique advantages, with KNN coming out on top. This research not only refines stress detection protocols but also sets the stage for future investigations into nuanced approaches for handling real-life stress datasets.

## Data availability

The data utilized in this paper has been obtained through a formal agreement with IMEC, OnePlanet Research Center, Netherlands. Any request for the data should be addressed to Prof. Chris Van Hoof, IMEC, OnePlanet Research Center, Netherlands.

